# Estimation of the epidemic properties of the 2019 novel coronavirus: A mathematical modeling study

**DOI:** 10.1101/2020.02.18.20024315

**Authors:** Jinghua Li, Yijing Wang, Stuart Gilmour, Mengying Wang, Daisuke Yoneoka, Ying Wang, Xinyi You, Jing Gu, Chun Hao, Liping Peng, Zhicheng Du, Dong Roman Xu, Yuantao Hao

**Affiliations:** School of Public Health, Sun Yat-Sen University, No.74, Zhongshan second road, Guangzhou, China; Sun Yat-sen University Global Health Institute, School of Public Health and Institute of State Governance, Sun Yat-sen University, Guangzhou, China; Graduate School of Public Health, St. Luke’s International University, Tokyo, Japan; School of Statistics, Jiangxi University of Finance and Economics, No. 169, East Shuanggang Road, Nanchang 330013, China; Health information center, Sun Yat-Sen University, Guangzhou, China

**Keywords:** COVID-19, novel coronavirus, epidemic planning, mathematical model, China, Wuhan

## Abstract

**Background:** The 2019 novel Coronavirus (COVID-19) emerged in Wuhan, China in December 2019 and has been spreading rapidly in China. Decisions about its pandemic threat and the appropriate level of public health response depend heavily on estimates of its basic reproduction number and assessments of interventions conducted in the early stages of the epidemic.

**Methods:** We conducted a mathematical modeling study using five independent methods to assess the basic reproduction number (R0) of COVID-19, using data on confirmed cases obtained from the China National Health Commission for the period 10^th^ January – 8^th^ February. We analyzed the data for the period before the closure of Wuhan city (10^th^ January – 23^rd^ January) and the post-closure period (23^rd^ January – 8^th^ February) and for the whole period, to assess both the epidemic risk of the virus and the effectiveness of the closure of Wuhan city on spread of COVID-19.

**Findings:** Before the closure of Wuhan city the basic reproduction number of COVID-19 was 4.38 (95% CI: 3.63 – 5.13), dropping to 3.41 (95% CI: 3.16 – 3.65) after the closure of Wuhan city. Over the entire epidemic period COVID-19 had a basic reproduction number of 3.39 (95% CI: 3.09 – 3.70), indicating it has a very high transmissibility.

**Interpretation:** COVID-19 is a highly transmissible virus with a very high risk of epidemic outbreak once it emerges in metropolitan areas. The closure of Wuhan city was effective in reducing the severity of the epidemic, but even after closure of the city and the subsequent expansion of that closure to other parts of Hubei the virus remained extremely infectious. Emergency planners in other cities should consider this high infectiousness when considering responses to this virus.

**Funding:** National Natural Science Foundation of China, China Medical Board, National Science and Technology Major Project of China

## Introduction

In December 2019 a novel coronavirus outbreak began in Wuhan, Hubei province, in China. As of 9^th^ February 37,558 cases of the virus had been confirmed globally, of which 37,251 were confirmed in China, with 813 deaths.^1^ On 30^th^ January the WHO declared the novel coronavirus (COVID-19) a public health emergency of international concern,^2^ and on 23^rd^ January the Hubei Provincial Government closed the city of Wuhan,^3^ followed by the closure of a wider network of cities in Hubei on 24^th^ January,^4^ to prevent its spread. Although the number of cases outside of China remains small, mathematical modeling has identified the risk of spread of the disease to population centres and transit hubs in other countries,^5^ with the possibility that the COVID-19 outbreak could become a global pandemic.

In the beginning stages of an epidemic, mathematical modeling is essential to understand the dynamics of the new disease, and to assess the organism’s infectiousness and rapidity of spread. This is primarily achieved by calculation of the basic reproduction number, denoted as *R*_0_, which measures the number of secondary cases that can be expected to be generated from a single case of the disease.^6^ Initial research from the first weeks of the COVID-19 outbreak estimated the basic reproduction number to be between 2.20 and 3.58, indicating large uncertainty in estimates of its infectiousness.^7,8^ Other unpublished estimates also placed the value of *R*_0_ within this range,^9^ with wide uncertainty.^5^ All of these estimates of the basic reproduction number were based on data to the end of January, and did not use a long series of data from the period after the closure of Wuhan city. The data series for these reports also did not include significant periods of time after the Chinese New Year (24^th^ January, 2020), when a large number of people return to their home towns from large cities, with the attendant risk of significant spread of the disease. Wuhan city has a population of 11 million people ^10^, but during the Chinese New Year as many as 5 million residents leave the city, and 70% of those who leave travel within Hubei province^11^, with the risk of significant spread of the disease within China, and especially across Hubei province, during the Chinese New Year period.

In this study we used data from the National Health Commission of the People’s Republic of China^12^ (NHC) for the period from the 10^th^ January to the 8^th^ February to estimate the basic reproduction number of COVID-19 using five mathematical modeling methods conducted independently. We used these modeling methods to estimate the basic reproduction number both before and after the closure of Wuhan city, and across the whole time period of the epidemic. Based on the results of these analyses we make recommendations for future control of the virus, and assess the future pandemic risk due to this new disease.

## Methods

Data was obtained from the NHC for the period 10^th^ January to 8^th^ February, 2020. The NHC is a cabinet-level executive department under the State Council of China which is responsible for public health, medical services and health emergencies in China. Data from before the 10^th^ January was not included in this analysis because cases identified before 10^th^ January were based on symptomatic diagnosis and no standardized testing method was available. Although the NHC provides information on suspected and confirmed cases, only data from confirmed cases was used in this analysis, to avoid confusion of routine seasonal influenza cases with nCoV.^13^

Confirmed cases were analysed using by applying four different estimation techniques, to allow for different assumptions about epidemic growth and the epidemiology of the disease:

- *Exponential growth (EG)*, which assumes an exponential growth curve to the virus and estimates the basic reproduction number from the Lotka-Euler equation^14^
- *Maximum likelihood method (ML)*, in which the likelihood of the cases is expressed directly in terms of *R*_0_ on the assumption of a simple SIR model structure^15^
- *Sequential Bayesian Method (SB)*, in which the posterior probability distribution of the basic reproduction number is estimated sequentially using the posterior at the previous time point as the new prior^16^
- *Time-dependent reproduction numbers (TD)*, in which the basic reproduction number at any time point is estimated as an average of accumulated estimates at previous time points^17^

These methods were implemented using the R0 package in R.^18^ All these models require no assumption about recovery time, but in some cases require an assumption about the generation time of the epidemic. All methods were applied to the data for the whole period (January 10^th^ to February 8^th^), to the period only before the closure (January 10^th^ to January 23^rd^), and to only the period after the closure of Wuhan city (January 23^rd^ to February 8^th^)

Because some of these methods are driven by a Suceptible-Infectious-Recovered (SIR) model, but an asymptomatic latent phase had been identified in the early progress of the disease, we also estimated the basic reproduction number using a standard Susceptible-Exposed-Infectious-Recovered (SEIR) model. An analytic expression for the basic reproduction number was obtained from the model using next generation matrices,^19^and predictions of cumulative cases were fitted to the data on national cases using maximum likelihood estimation to identify the optimal value of *R*_0_.^20^A Metropolis-Hastings Monte Carlo sampling method^21^ was used to estimate a credible interval for the basic reproduction number. In this model the latent phase was fixed to last 5.2 days,^8^ and the recovery period was fixed at 7 days. Although the biological recovery period of the disease has not been clearly established, the period from onset of symptoms to isolation in specialist hospitals was assumed to be 7 days, and the recovery compartment of the SEIR model acts as a proxy for isolation in these models. All MH estimates used 20,000 Monte Carlo Iterations with a burn-in period of 5,000 iterations and a normally distributed proposal distribution.

Because all five modeling methods use different assumptions and tools and are likely to produce widely varying estimates of the basic reproduction number based on different aspects of the epidemic process, we combined all five estimates to produce an overall value for the basic reproduction number. We calculated a weighted average of the five basic reproduction numbers by calculating weights from a Poisson loss function,^22^ which is similar to a Poisson likelihood but which was not used for estimation of parameters in any of the models. We also estimated a weighted standard error from the models. Where standard errors do not overlap point estimates in the pre- and post-closure periods, we conclude there is a significant difference in the epidemic process between the periods. Finally, we estimated the predicted epidemic trend from all models in each period, and plotted these against the observed number of cases for each period.

Mathematical details of the models are presented in the supplementary appendix. All results are presented as values of the basic reproduction number with its 95% confidence interval. For the MH method the inter-quartile range of the posterior distribution of *R*_0_ is presented.

### Role of the funding source

The funders had no involvement in study design, data collection, analysis or interpretation of the data, had no influence on the writing of the report or the decision to submit for publication.

## Results

By February 8^th^ there were 37,198 confirmed cases nationally, with 27,100 of these cases in Hubei (72.9% of all cases), and 14,982 in Wuhan (40.2% of all cases). All models applied to these data estimated the basic reproduction number effectively. Basic reproduction numbers for all fives methods for the entire time period, the pre-closure period and the post-closure period, are shown in Table 1. The best-fitting method in the entire period was the method based on time-dependent reproduction numbers, while the pre-closure and post-closure period were best fitted by the exponential growth model.

**Table 1:**
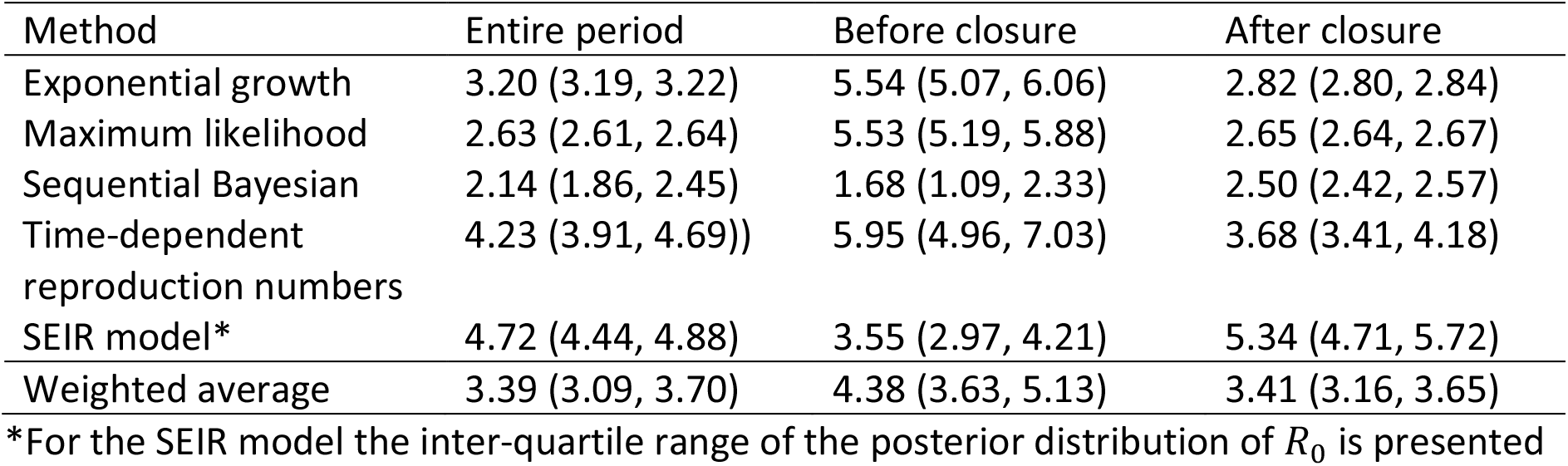
Estimates of the basic reproduction number for 5 methods by epidemic period, with weighted average

The weighted average estimate of the basic reproduction number shows that the epidemic slowed down after the closure of Wuhan city, dropping from 4.38 (95% CI 3.63 – 5.13) before the closure to 3.41 (95% CI 3.16 – 3.65) after. The 95% confidence intervals for the exponential growth estimate of post-closure *R*_0_ do not overlap the point estimate for the pre-closure period, indicating that there was a significant reduction in the basic reproduction number after the closure of Wuhan. Figure 1 shows the model predictions from all five models plotted against the observed cases for the pre-closure period (top left panel), post-closure period (top right panel) and entire period (bottom panel). A similar figure, with only the best-fitting model shown, is given in Supplementary Figure S2.

**Figure 1:**
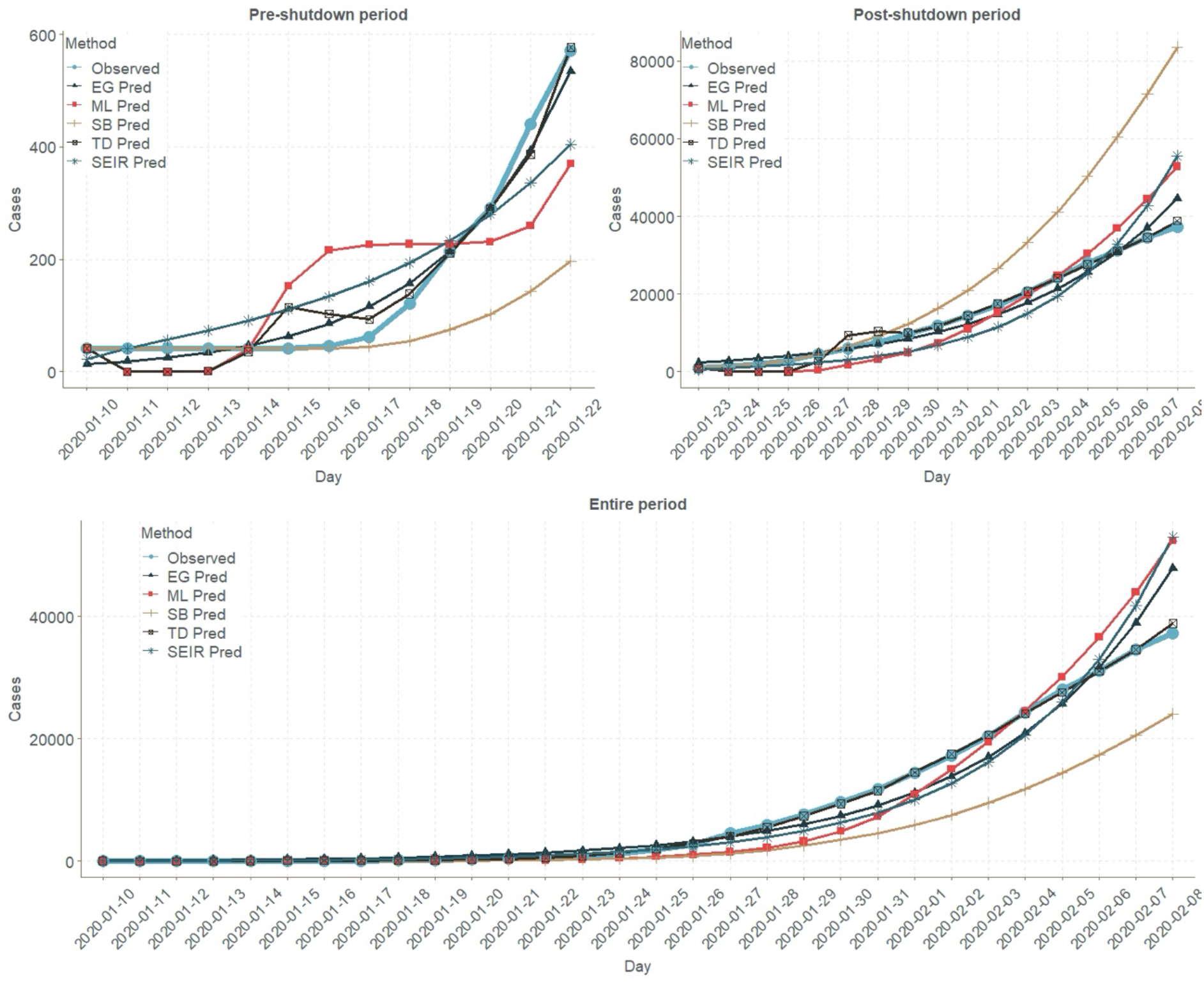
Predicted epidemic curves for the pre-closure, post-closure, and entire epidemic period estimated from five models, with observed values

From figure 1 it is clear that models that estimated low values for the basic reproduction number in the pre-closure period or the entire period, such as the Sequential Bayesian model, produced very poor predictions that under-estimated the epidemic, and the best-fitting models were those that identified basic reproduction numbers over 4 in the pre-closure period, and over 3 in the entire epidemic period.

## Discussion

This study estimated the basic reproduction number, *R*_0_, for the 2019 novel coronavirus using confirmed cases from 10^th^ January – 8^th^ February. We applied five methods to estimate *R*_0_ in order to ensure that our estimate was robust to differences in assumptions about epidemic processes, differences in assumed underlying parameters, and about the nature of the dynamics of the affected population. We estimated the basic reproduction number for the whole period to be 3.39 (95% CI: 3.09 – 3.70), a very high number indicative of a very fast rate of spread. We also estimated the basic reproduction number separately for the pre-closure period, finding that in the first 13 days of available high-quality data on the epidemic that the basic reproduction number was 4.38 (95% CI: 3.63 – 5.13), a very high number indicative of a highly infectious disease. Compared to this, we calculated the post-closure value of *R*_0_ to be 3.41 (95% CI: 3.16 – 3.65), still a very high number but significantly lower than that observed in the earlier days of the epidemic. This lower basic reproduction number, and the recent apparent reduction in numbers of new infections, justifies the decision to close Wuhan city, and also shows the potentially high impact of self-quarantine and voluntary exclusion methods. A separate study (not published) conducted by one of the study authors found that 25% of students in Guangdong did not leave their home in the Chinese New Year period, 90% increased their handwashing frequency, and over 80% used a mask when moving in public places. These voluntary measures, combined with the closure of Wuhan city, may have averted the spread of this disease and reduced its ability to reproduce through social changes. The reduction in infectiousness is particularly striking given the huge movement of people that typically occurs during Chinese New Year, and the risk of exposure in public transport and family gatherings at this time.

A striking feature of our analysis is the very high value of the basic reproduction number we identified in the period of time up to the closure of Wuhan city. Three of our modeling methods – including the best-fitting method based on a Poisson loss function – identified a value of *R*_0_ greater than 5, with some possibility of a value over 6. Basic reproduction numbers in the 5-7 range are consistent with extremely contagious diseases such as mumps and smallpox, and indicate a disease with a very high risk of becoming a global pandemic. This finding has significant implications for cities like Singapore, Japan and London which are beginning to experience the first signs of spread of the disease without importation. In light of the epidemic threat identified here, these cities should consider implementing more aggressive prevention policies as necessary, while respecting human rights and the dignity of affected individuals and of those who might be disadvantaged by stricter quarantine and control mechanisms.

Previous studies^5,7-9^ found lower values for the basic reproduction number. This variation may have arisen for two reasons. First, the empirical data that previous studies used were collected before 25^th^ Jan, 2020. Testing protocols and diagnostic tools changed during the early period of the study^23^, and the number of diagnosed cases collected before 15^th^ January were considered underestimated and less reliable. This would flatten the epidemic curve in early studies, and the estimation of R0 based on these data may be underestimated and have larger confidence intervals. Second, previous studies only estimated the R0 based on a single method, and these estimates may have been affected by the implicit assumptions in these models. For example, a previous paper using the assumption of exponential growth found a value of *R*_0_ of 2.68 (95% Credible interval 2.47 – 2.86)^5^ using an SEIR model with Metropolis-Hastings MCMC estimates of uncertainty, but our modeling has shown that this method likely underestimated the basic reproduction number during the pre-closure period. Our model avoids the limitations of specific modeling choices by combining several methods with a Poisson Loss weight, using the most current and accurate case diagnosis. Through this approach we calculate a more robust estimate than previous studies, and find a higher value of *R*_0_. Ours is also the first study to compare the pre- and post-closure periods in the data, and thus the first study to make a judgment about the effectiveness of this strategy. Given the high risk of epidemic from COVID-19, it is important to assess the value of this strategy before the disease takes hold in another global city.

This study has several limitations. It was based on confirmed cases, and by excluding suspected cases or mild cases may have under-estimated the rate of spread of the disease. We did not estimate the values of the parameters defining the transition rate from exposed to infectious, or infected to recovered, but fixed them at previously published values. This was a necessary decision because the clinical features of the disease are not yet fully understood, and may affect estimates. However, our intuition after fitting these models is that the maximum likelihood estimate of the force of infection naturally adjusts to fit the value of the recovery rate, and produces a broadly similar value of the basic reproduction number as a result. Furthermore, to adjust for the still-arbitrary nature of these estimates of key parameters, we used some methods that do not depend on any assumptions about these aspects of the disease process. Another limitation of this study is the uncertainty introduced by the use of multiple modeling methods, which we combined with a weighted average. However, we believe that by presenting varying methods with different assumptions along with a weighted average, we enable researchers and policy-makers to make judgements about the dangers of the epidemic without relying on any particular set of assumptions about a disease that is not yet well understood.

Our results show that the 2019 novel Coronavirus is an extremely rapidly spreading and dangerous infectious disease, with the potential to infect a very large proportion of the population very rapidly if not contained. Extreme epidemic prevention measures, including city closures and wide-scale self-quarantine, may be necessary simply to reduce the pace of the epidemic, and even these extreme measures may not be sufficient to prevent pandemic. City officials, public health planners, policy-makers and governments in countries that are beginning to see the spread of this disease domestically need to act quickly, effectively and decisively, as the government of China did, to prevent a serious global pandemic.

## Data Availability

All data used in this manuscript was obtained from the China National Health Commission and is available from them on request.

## Author contributions

YH and JL conceived of the study. JL, SG, YJW, MW, DY, XY and YW conducted data analysis. SG, YJW, MW, DY and JL wrote the first draft of the paper. JL, JG, CH, ZD and LP collected data. All authors offered scientific input and edited all drafts of the paper.

## Declaration of interests

The authors have no conflict of interests to declare.

## Acknowledgments

JL is supported by the National Natural Science Foundation of China (grant ID 81803334) and China Medical Board (grant ID 18-301). JG is supported by the National Natural Science Foundation of China (grant ID 71774178). CH is supported by the National Natural Science Foundation of China (grant ID 71974212). YH is supported by the National Natural Science Foundation of China (grant ID 81973150) and the National Science and Technology Major Project of China (grant ID 2018ZX10715004).

## Notes

### Competing Interest Statement

The authors have declared no competing interest.

### Funding Statement

This project was partially funded by:
The National Natural Science Foundation of China, China Medical Board, National Science and Technology Major Project of China

